# Comparing Esri ArcGIS and SAS Geocoding Approaches: Test case with 3,238 Wisconsin addresses

**DOI:** 10.1101/2025.01.23.25321040

**Authors:** Hannah K. Johnson, John M. Hampton, Natalia Arroyo, Amy Schultz, Ronald E. Gangnon, Kristen M. Malecki, Amy Trentham-Dietz

## Abstract

This report describes a comparison of two geocoding methods used by the Cohorts for Environmental Exposures and Cancer Risks (CEECR) consortium: ArcGIS Geocoding by Esri and the SAS GEOCODE Procedure. The goal of this report is to determine the comparability of data sets that employ different approaches for linking survey data with spatial surrogates of exposure to environmental and socioeconomic factors. ArcGIS and SAS GEOCODE were selected as two platforms for this comparison because both programs are being used by one or more CEECR cohort study teams and they can be used locally offline. The latter minimizes confidentiality issues related to online data linkages. Residential addresses from 3,238 Wisconsin residents in the *Cancer & COVID Study* and the *Wisconsin in situ Cohort* (WISC) were geocoded and linked to eight different publicly available datasets of environmental and socioeconomic factors at various geographic scales using both geocoding platforms. Since the two analytic platforms vary in geocoding approaches, the validity and accuracy of both platforms were compared to examine differences when assigning surrogate measurements of exposure based on spatial locations. ArcGIS offered a higher specificity for matched addresses with slightly more latitude/longitude point and street matches (97.7%) than SAS (95.9%), with the remainder matching at the zip code level. The two geocoding platforms showed high concordance in assignment at the county (99.6%), census tract (96.5%), and census block group (94.7%). As a result, the correlations based on census tracts and block groups were very strong for linked exposure measures of socioeconomic status, environmental justice, urban/rural residence, air pollution, proximity to industrial sites, and cancer risk (all intraclass correlation coefficients ≥98%). Slightly lower concordance was observed for point source linkages (intraclass correlation coefficients 96-97%). Approximately ~4% of addresses were mis-matched largely in rural areas where census areas are larger and accurate geocoding base-layers are less widely available than in urban areas. For researchers that are already utilizing SAS, the GEOCODE procedure can be a logical choice as it is included in base SAS software and does not require an additional cost. However, SAS and ArcGIS provide similar options for the vast majority of study address locations.

## INTRODUCTION

The goal of this report is to determine the comparability of data sets that employ different approaches for linking survey data with spatial surrogates of exposure to environmental and socioeconomic factors. Addresses of study participants are often geocoded in research studies to link with external databases to advance exposure assessment for environmental and social exposures. Numerous social and economic data are available from the census and other sources to describe neighborhood characteristics including social determinants of health. Similarly, many environmental indicators, such as traffic density or industries reporting to the Toxic Release Inventory, can be used to estimate potential surrogate measurements of chemical and non-chemical exposures, such as air pollution. In this report, we describe concordance in the two geocoding methods for assigning addresses to the same geographic unit (e.g., latitude/longitude, census tract, zip code, etc.) and for linkage to spatial measures of social and environmental exposures available in existing online databases.

CEECR researchers are currently using two geocoding methods: ArcGIS Geocoding by Esri (Redlands, CA) and the GEOCODE procedure in SAS (Cary, NC: SAS Institute Inc). These two geocoding platforms perform their geocoding matches for input addresses in different ways. Thus, comparison of the two geocoding methods is important for intended collaborative geospatial research in the CEECR consortium. We anticipate higher mismatch rates and disagreement between geocoding outputs from the two geocoding methods for rural addresses and addresses in areas with more residents with lower socioeconomic status (SES, e.g., lower annual household income) (1, 2). We also expect that any linkages will be dependent upon the size of the geographic unit since smaller units such as census tracts may link less consistently with other data such as counties. Thus, data linkages that require larger geographic areas may result in fewer observed differences between the two geocoding methods.

## OBJECTIVE

We sought to conduct a reliability study using mailing addresses reported by participants from two Wisconsin-based epidemiologic cancer studies. Specific objectives include:

- Assess the quality of match rates for the two geocoding methods according to different geographic scales (latitude/longitude, street, zip code)
- Quantify the level of agreement between the two geocoder latitudes/longitudes in distance proximity and concordance of geographic units
- Quantify level of agreement when linked to area-level social and environmental datasets
- Identify sources of mismatches and disagreements between the two geocoding methods

## METHODS

### Mailing address sources and preparation for geocoding

Addresses for this comparative analysis came from two studies conducted in Wisconsin: one titled *The impact of coronavirus disease 2019 on Wisconsin cancer patients* (i.e., *Cancer & COVID* [3]) and the other titled: *The Wisconsin in situ Cohort* (*WISC* [4]). Additional information on these studies and initial processing prior to geocoding is summarized in the table below (**Table 1**). This project was approved by the University of Wisconsin Health Sciences Institutional Review Board.

**Table 1.**
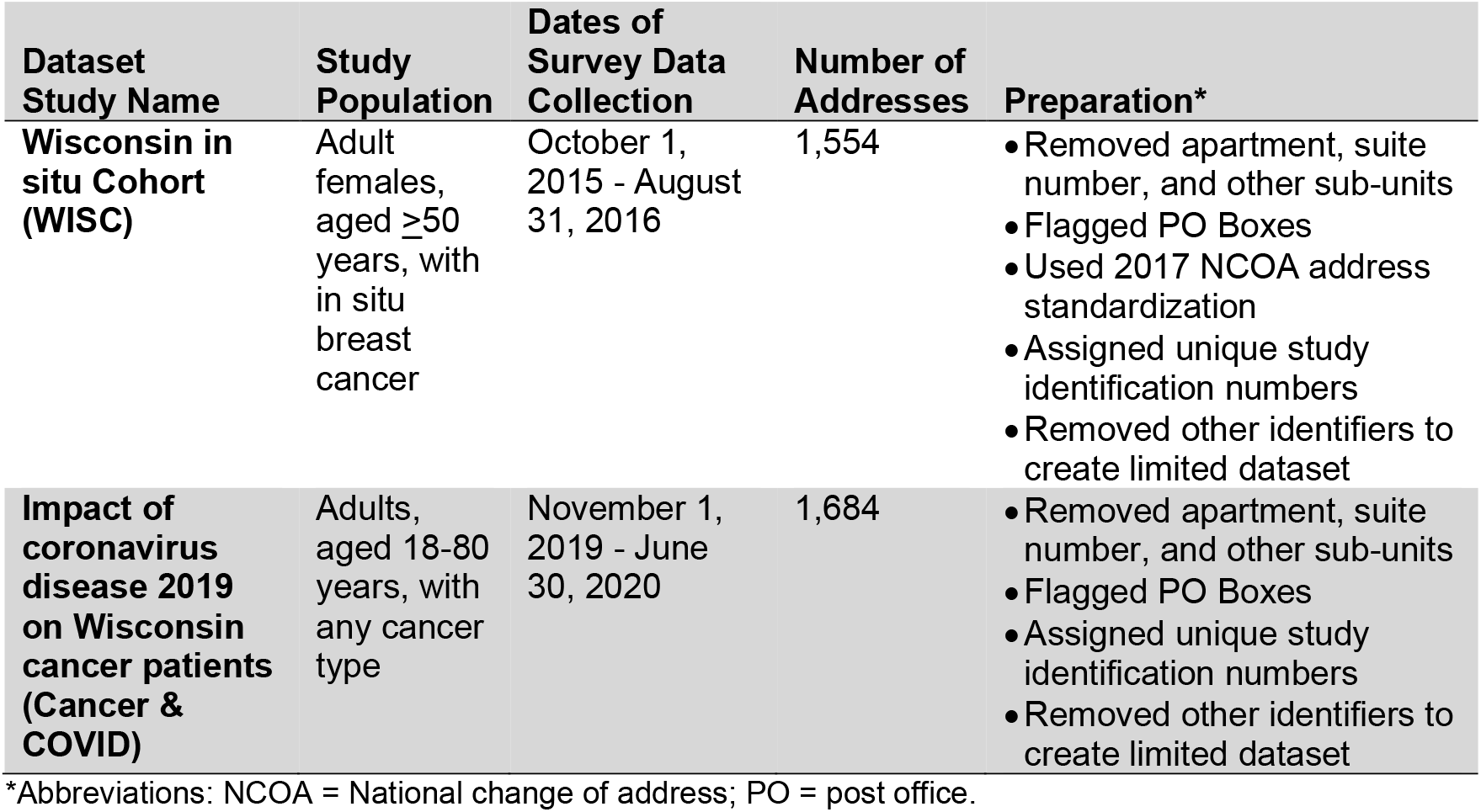
Study sources for addresses and processing for geocoding.

Each study participant had one mailing address geocoded. For *Cancer & COVID* (a cross-sectional study with a single survey), the enrollment address was used; for *WISC* (a cohort study with repeated surveys), the last known address was geocoded, and former addresses were not examined. Participants in both studies were adults living in Wisconsin, survivors of cancer, and completed study questionnaires in English. *WISC* study participants were initially identified from the state cancer registry in 2015 and 2016, while *Cancer & COVID* participants were identified from the cancer registry operated at the University of Wisconsin Carbone Cancer Center for patients who received care at one of their cancer clinics in 2019. Most study participants were female, over 60 years of age, non-Hispanic White, and had attended college (**Supplementary Table 1** in the **Appendix**).

To prepare for geocoding, addresses were assigned unique study identification numbers and standardized in SAS (**Table 1**). Identifiers other than participant address were removed from the datasets for the geocoding comparison. Addresses outside of Wisconsin were removed from the dataset. *WISC* addresses, but not addresses for the *Cancer & Covid* study, were standardized using the 2017 National Change of Address (NOCA) service by the US Postal Service. Apartment, suite numbers, and other sub-units were removed from the original mailing addresses so they would not interfere with geocoding. In addition, a new variable was created in SAS that flagged PO box addresses in the dataset.

### Overview of geocoding methods

Two geocoding methods were examined in this report. A summary of key features of the different applications and their methods for point level geocoding (creating latitudes and longitudes) can be found below.

#### ArcGIS (Redlands, CA: Esri): ArcGIS StreetMap Premium Offline Geocoder

- Procedure - The prepared address file for each study was converted from Excel (.xlsx) to .csv for upload into ArcGIS Pro Desktop version 3.2. Wisconsin shapefile census data from 2018 and 2021 was added using TIGER/Line files for census tract, block group, and county. Shapefiles from 2018 census data were used for the *WISC* dataset and shapefiles from 2021 census data were used for *Cancer & COVID* (5). The ArcGIS spatial join tool was used to link the geocoded addresses (points) to geographic units (spatial polygons from TIGER/Line Shapefiles for county, tract, and block group). Participant addresses were geocoded to point addresses using the locator for USA as the study only included Wisconsin residents. Publicly available datasets describing social and environmental exposures or cancer risk also were converted, if necessary, to the .csv format for upload into ArcGIS. When geocoded to a point address, each participant address was matched to an address in the locator file, which included a latitude and a longitude for the point location of that matched address. ArcGIS assigns locations based on “exact centroids” of buildings although the source of the location information for buildings (including residential homes) is unclear. If location information was not available, ArcGIS used the same process as SAS to assign a location, in other words, by interpolating the location of an address assuming equally spaced addresses along a street segment.
- Scoring – ArcGIS assigned a match score of the geocode to each study participant’s inputted address. The score ranged from 0 to 100, where 100 indicates a perfect match between the inputted address and the matched address from the ArcGIS locator file. The match score is based on how well the address identified in the reference dataset during geocoding matches to the inputted participant address. A 0 match score indicates that no such address was found in the reference dataset (6). The scores are based on a weighted numbering system of matching characters in each of the address element areas; as more characters match, the score increases. Unlike SAS Scoring, missing elements do not penalize the match score; instead, missing elements do not contribute to the total (6). More information on how the match score is calculated in ArcGIS can be found in this thread on StackExchange that asks “How are geocoding scores calculated in ArcGIS?” (6).
- Confidentiality - To comply with HIPAA requirements and maintain participant confidentiality, the geocoding process was conducted in a secure offline environment using ArcGIS StreetMap Premium software (ArcGIS Pro Desktop Version 3.2). Only addresses were geocoded and no other personal identifying information was used to geocode addresses and link to exposure measures. Linkages were done after processing for geocoding was complete.

#### SAS PROC GEOCODE: SAS version 9.4 Windowing Environment

- Procedure - The SAS-specific 2018 and 2021 TIGER/Line files used in the GEOCODE procedure were downloaded from the SAS Maps and Geocoding database (5). The GEOCODE procedure attempts to match the address inputted to an address in the lookup data set from SAS using either STREET or PLUS4 geocoding methods (8). If no match is found at these levels, then it will search for a five-digit zip code match for the input address (8). For addresses matching at the street level, SAS PROC GEOCODE (using method=street) links the street address to latitude and longitude coordinates, state, county, tract, and block group (8). For addresses that do not match at the street level but do match at the 5-digit zip code level, SAS PROC GEOCODE links the address to zip centroid latitude and longitude coordinates but does not include county, tract, and block group. To add the county, tract, and block group for the zip centroid geocoded address, the GINSIDE SAS procedure was utilized. Note that SAS does not geocode an address to a specific house number or building. Instead, SAS PROC GEOCODE approximates the position of a particular address on a street by assuming that house numbers are an equal distance apart.
- Scoring – For all matches (street-level or zip code-level), the GEOCODE procedure produces a match score, which is a numeric value indicating the relative accuracy of the match. The score is based on several components of the address match and each token in the _NOTES_ value has an associated value, with the sum of these values making up the value of the _SCORE_ variable. The scoring algorithm adds each component score and subtracts identifiable differences between matches; higher scores are “better”. There is not a perfect score; the score depends on what components of the address are entered and which ones matched or did not match. Of note, in SAS 9.4M6, abbreviated address parts that result in a match are scored lower than fully named, similar address parts (e.g., LN versus LANE).
- Confidentiality - To comply with HIPAA requirements and maintain participant confidentiality, the geocoding process was conducted in a secure offline environment using SAS version 9.4 in the windowing environment.

### Linkages to external datasets

Geocode outputs (e.g., latitude and longitudes) from the SAS and ArcGIS geocoders for the *WISC* and *Cancer & COVID* studies were compared for match accuracy and then linked to several publicly available geographic area-level datasets. Exposure assessment linkages with social and environmental data were compared at multiple scales. A summary of data sources used in the comparisons by level of assessment is provided below and in **Supplementary Table 2** in the **Appendix**.

#### Using County

- The Rural-Urban Continuum Codes (RUCCs) were linked to the study addresses at the county level. RUCC codes are based on population size and proximity to a metropolitan area with codes 1 to 3 representing more populous metropolitan counties and codes 4 to 9 representing nonmetropolitan and less populous counties (9).

#### Using Census Tract

- The Yost Index for the years 2014 – 2018 was linked to the study addresses based on census tract (8). The Yost Index is a measure of socioeconomic status across geographic areas that combines educational level, income, housing, and employment information.
- The Yost Index is a percentile score from 1 (most affluent) to 100 (most deprived). Authors of the Yost Index used five-year ranges to balance granularity of data from the American Community Survey while also reducing file size for download (11).
The Environmental Justice (EJ) Index for 2022 ranks each census tract on 36 environmental, social, and health factors and groups them into three overarching modules (Social Vulnerability, Environmental Burden, and Health Vulnerability) and ten different domains (12). The EJ Index Social-Environmental Ranking (SER) is calculated by combining rankings from the Environmental Burden Module and the Social Vulnerability Module, but not from the Health Vulnerability Module. The SER has a 0-1 range.
- Rural-Urban Community Area Code (RUCA) is a measure of urbanization, population density, and daily commuting (13). RUCA scores range from 1-10 and are divided into 4 groups with 1 to 3 representing metropolitan areas, 4 to 6 micropolitan areas, 7 to 9 small towns, and 10 rural areas. The RUCA 2010 values were revised in 2019.
- The Air Toxics Screening Assessment (AirToxScreen) is Environmental Protection Agency’s screening tool that provides communities with information about health risks from air toxics. Specifically, AirToxScreen estimates the cancer risks from breathing air toxics over many years. Values for the National Cancer Risk by Pollutant for 2019 were obtained for this project (13). The National Cancer Risk by Pollutant dataset includes a lifetime cancer estimate for geographic entities throughout the United States. National Cancer Risk is expressed as “N” – in – 1 million people where “N” is the number of people for every 1 million people who are continuously exposed to a certain level of a pollutant over 70 years that may develop cancer (15). Any value greater than 100 in 1 million for a census tract (1 in 10,000) was intended to represent an elevated cancer risk in that tract (16). Air toxins included in the dataset are known to cause or suspected to cause cancer or other serious health effects (15).

#### Using Block Group

- The Yost Index (described above) was linked to addresses at the block group level in addition to the census tract level (10).

#### Using Small-Scale Grids

- Particulate matter less than 2.5 µm (PM2.5) grided downscaled air pollution data for 2020 was linked based on latitude/longitude coordinates of the geocoded output files to the latitude/longitude coordinates for nearest grid centroid point (17). Using Microsoft Access, the downloadable spreadsheet was filtered between −86 and −93 decimal degrees and 42 and 48 decimal degrees to make a square of grid points for Wisconsin. After the file was downloaded, annual means for PM2.5 daily values were calculated in SAS using PROC MEAN. This data was then used in ArcGIS. The 12km grid centroid points were overlayed on top of the ArcGIS basemap with the latitude/longitude points for the datasets and then the nearest grid point was determined using the ‘Near’ function in ArcGIS. Similarly, in SAS, the nearest grid point was identified by joining the 12km grid centroid point file with the geocoded addresses file, calculating the distance between an individual address to each grid point using the GEODIST SAS function, then reducing the dataset to the nearest distance for each address.

#### Using Point Source (latitude/longitude)

- Toxic Release Inventory (TRI) data for Wisconsin in 2022 was downloaded using the EPA TRI Explorer tool (18). Distance to the nearest TRI facility was calculated using the ‘Near’ function in ArcGIS or using the GEODIST function in SAS.

### Statistical analysis

The two geocoding methods were compared in four ways corresponding to the items listed in the Objective section above:

- *Assess the quality of match rates for the two geocoding methods according to different geographic scales*. For the purposes of this assessment, a ‘match’ refers to the individual geocoders assigning a latitude/longitude point to the input addresses. ‘Specificity’ refers to the address match type a geocoder assigned to an input address. A point address, for example, is more specific to a geographic location than a zip code. When linking to environmental exposure datasets using smaller geographic units like point, block group, or tract, specificity is particularly important as it can impact exposure characterizations (19, 20).
- *Quantify the level of agreement between the two geocoder latitudes/longitudes in distance proximity and concordance of geographic units*. Distance was measured in miles. Four geographic units were used including census block group, census tract, county, and state.
- *Quantify the level of agreement when linked to area-level social and environmental datasets*. Concordance between exposure levels assigned by the two geocoding methods was measured using intraclass correlation coefficients with 95% confidence intervals (CI).
- *Identify sources of mismatches and disagreements between the two geocoding methods*. ‘Discordance’ refers to differences observed between the two geocoding outputs at the point, tract, block group, county, or state levels. P-values from chi-square tests were calculated for the test of whether the distance between the geocoded locations was distributed in categories with the same frequency according to rural residence (measured by RUCC), Yost Index, and geographic unit for the match (latitude/longitude coordinate, street, or zip code).

## RESULTS

### Quality of match rates

All addresses were assigned a geocode; in other words, none of the addresses failed to match in either ArcGIS or SAS. The median match score assigned to the geocoded address for the SAS geocoder was 75 (range, 2-105) (**Table 2**). The median match score for the ArcGIS geocoded addresses was 100, which represented perfect matches to point addresses. SAS scores tended to vary more than ArcGIS scores. The minimum score in ArcGIS of 84.4 was a match to a postal code point in the reference dataset, which was due to a missing street name and number in the input data.

**Table 2.** Descriptive statistics of geocoding scores provided by the two geocoding methods for 3,238 Wisconsin addresses.

| Statistics | SAS | ArcGIS |
| --- | --- | --- |
| Minimum | 2 | 84.4 |
| 25th % | 75 | 100 |
| Median | 75 | 100 |
| 75th % | 75 | 100 |
| Maximum | 105 | 100 |
| Mean | 72.9 | 99.8 |
| Standard deviation | 16.4 | 1.3 |

About 95% of addresses were geocoded to the smallest geographic unit available in both geocoders, equal to the street level in SAS and latitude/longitude coordinate in ArcGIS (**Table 3**).

**Table 3.**
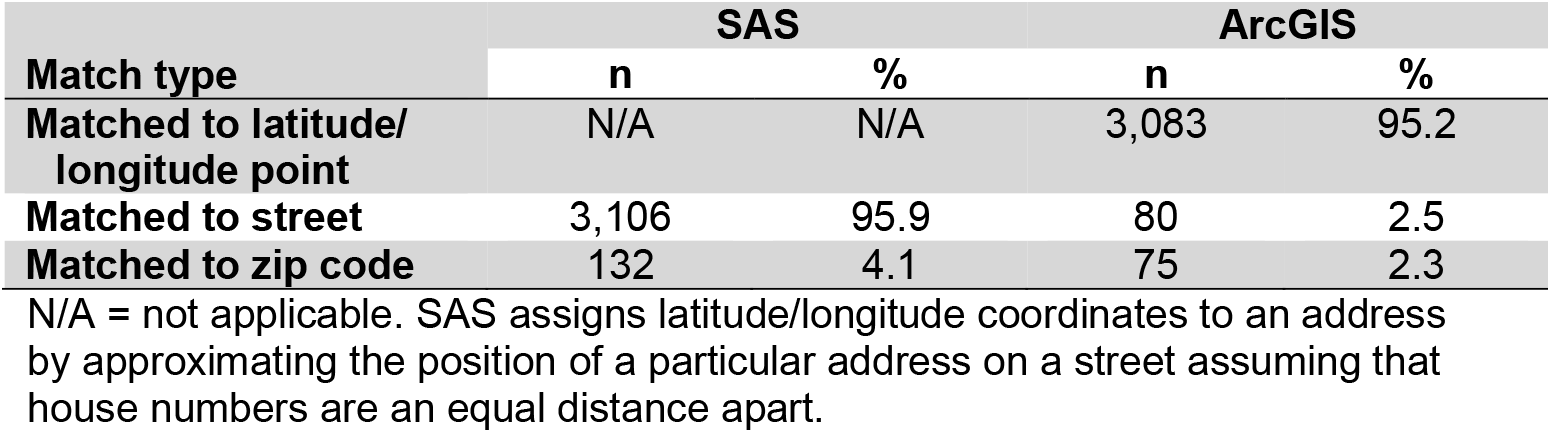
Match type specificity for the two geocoding methods based on 3,238 Wisconsin addresses.

### Geocoder concordance

Since each geocoding method used different methods to calculate match scores, we did not compare the match scores and instead calculated the distance between the geocoded locations. The median distance between geocoded latitude/longitude points from the two geocoders provided a median value of 0.03 miles with a range of 0 to 16.7 miles (**Table 4**, latitude/longitude point is approximated in SAS based on an estimated position on a street).

**Table 4.** Descriptive statistics of distance between geocoded latitude/longitude coordinates for 3,238 Wisconsin addresses.

| Statistics | Distance (miles) |
| --- | --- |
| Minimum | 0.00 |
| 25th % | 0.02 |
| Median | 0.03 |
| 75th % | 0.07 |
| Maximum | 16.7 |
| Mean | 0.2 |

Latitude/longitude coordinates from the two geocoders were within 0.5 miles of each other for 93.9% of the addresses with 94.7% assigned to the same census block group (**Table 5**).

**Table 5.**
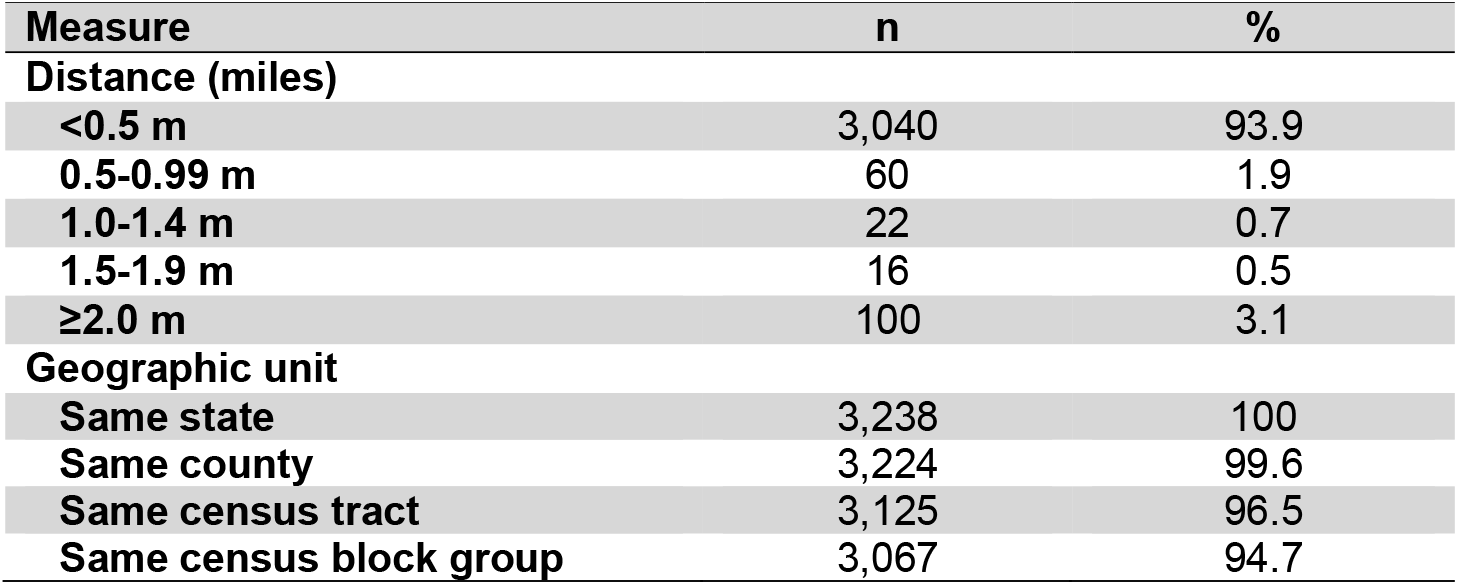
Concordance of distance and geographic units between the two geocoders for 3,238 Wisconsin addresses.

| Measure | n | % |
| --- | --- | --- |
| **Distance (miles)** |  |  |
| <0.5 m | 3,040 | 93.9 |
| 0.5-0.99 m | 60 | 1.9 |
| 1.0-1.4 m | 22 | 0.7 |
| 1.5-1.9 m | 16 | 0.5 |
| ≥2.0 m | 100 | 3.1 |
| **Geographic unit** |  |  |
| Same state | 3,238 | 100 |
| Same county | 3,224 | 99.6 |
| Same census tract | 3,125 | 96.5 |
| Same census block group | 3,067 | 94.7 |

### Agreement of environmental and cancer measures from linked datasets

Comparing area-level measures corresponding to geocoded points resulted in uniformly high intraclass correlation coefficient values (0.98-1.00) (**Table 6**; **Appendix 1, Table 2**). The correlation in area-level measures assigned by the two geocoders was slightly higher for the measurement that was linked based on county (RUCC) as compared with measures based on smaller geographic units (Yost, EJ Index, RUCA) (**Table 6**).

**Table 6.** Correlation between geographic area-level measures for 3,238 Wisconsin addresses.

| Measure | Geographic Unit | Intraclass Correlation Coefficient | 95% CI |
| --- | --- | --- | --- |
| Yost Index percentile | Block group | 0.98 | 0.979-0.982 |
| Yost Index percentile* | Census tract | 0.98 | 0.979-0.982 |
| Environmental Justice (EJ) Index Social-Environmental Ranking* | Census tract | 0.98 | 0.979-0.981 |
| Rural-Urban Commuting Area Code (RUCA) | Census tract | 0.99 | 0.990-0.992 |
| Air Tox Screen Total Cancer Risk (per million) | Census tract | 0.98 | 0.984-0.986 |
| Rural-Urban Continuum Code (RUCC) | County | 1.00 | 0.998-0.998 |

\*3,237 addresses due to 1 missing value

The two geocoders also showed high concordance for assigning addresses to the closest Toxic Release Inventory facility and the closest 12km grid with PM2.5 measures (>96% concordance) (**Table 7**).

**Table 7.** Concordance of coordinates for geographic-area level measures for 3,238 Wisconsin addresses.

| Measure | Number of Addresses | % |
| --- | --- | --- |
| Toxic Release Inventory facility | 3,122 | 96.4 |
| PM2.5 Grid | 3,136 | 96.9 |

### Sources of discordance between geocoded coordinates

The two geocoders were more likely to assign addresses to distances at least 1 mile apart if the addresses were located in a non-metropolitan county and if either of the geocoders matched the addresses at the zip code level rather than point or street level (**Table 8**). Geocoded addresses were equally likely to be at least 1 mile apart based on a lower or higher Yost Index social disadvantage value.

**Table 8.** Distance between geocoded point locations by SAS and ArcGIS according to key factors for 3,238 Wisconsin addresses.

| **Factor** | **Points <1 mile apart** | **Points ≥1 mile apart** | **P-value from Chi-square Test** |
| --- | --- | --- | --- |
| **RUCC** |  |  | <.0001 |
| Metropolitan | 2,312 | 78 (3.3%) |  |
| Nonmetropolitan | 787 | 60 (7.1%) |  |
| **Yost Index** |  |  | 0.233 |
| <50 (lower disadvantage) | 2,191 | 91 (4.0%) |  |
| ≥50 (greater disadvantage) | 909 | 47 (4.9%) |  |
| **SAS matches\*** |  |  | <.0001 |
| by street | 3,042 | 64 (2.1%) |  |
| by zip code | 58 | 74 (56.1%) |  |
| **ArcGIS matches†** |  |  | <.0001 |
| by point | 2,995 | 88 (2.9%) |  |
| by street | 61 | 19 (23.8%) |  |
| by zip code | 44 | 31 (41.3%) |  |

\*ArcGIS matches at all geographic units
†SAS matches at all geographic units

## CONCLUSIONS

Two geocoding methods - ArcGIS and SAS Geocode - were of interest by the CEECR investigators. Geocoding outputs were similar across both geocoding software methods, and they did not appear to substantively impact linkages to external datasets. While numerous geocoding applications are now available, these two were selected because they can be installed on secure desktop servers and used without connecting to a cloud-based server, thereby helping to preserve confidentiality.

For this dataset consisting entirely of Wisconsin addresses, ArcGIS offered a higher specificity for matched addresses with slightly more latitude/longitude point and street matches (97.7%) than SAS (95.9%), with the remainder matching at the zip code level. As found in this analysis, nonmetropolitan or rural addresses have been shown to produce more geocoding error than metropolitan or urban addresses; some states can have more accurate geocodes than others (1, 21). Researchers with a large proportion of study participants living in rural areas may want to select ArcGIS for geocoding. However, the source of data used by ArcGIS to assign latitude/longitude coordinates to buildings is not well described, thus the modestly improved specificity may be artifactual. Without an analysis with true location data available for buildings with their assigned addresses, it is difficult to know whether ArcGIS’s data on buildings is more accurate than SAS’s estimates based on equally spaced addresses along a street.

For data linkages to environmental exposure or other area-based measures, use of either geocoding software tool is likely to be concordant for most addresses. SAS PROC GEOCODE does not require an additional cost for researchers already using SAS. Use of either SAS or ArcGIS geocoder should not significantly impact collaborative projects within research consortia like CEECR that rely on geographic linkages. Both geocoders can be used offline, which provides greater protections against losses of confidentiality for study participants.

A geocode based on a zip code from any software tool is typically considered a poor-quality match and should not be used for geographic linkages that are smaller than a state. These are the same addresses where SAS and ArcGIS were most likely to assign latitude/longitude locations more than a mile part. Conversely, a geocoded location based on a street address is likely to consistently assign the same census tract regardless of geocoder software, with high confidence for matching to any geographic unit larger than census tract.

This study had strengths and limitations, especially related to generalizability of the findings. Study addresses were selected for this geocoder comparison due to their convenience (large number, already prepared for geocoding), statewide distribution, and the recency of the studies. All geocoded addresses were in Wisconsin. Study participants were cancer survivors with one study limited to women with early-stage breast cancer. Most participants in both studies were over 60 years of age. Approximately 30% of Wisconsin residents live in rural areas as compared to 19% of US residents. Most study participants also reported their race as White, and populations are known to have different patterns of residential location in the US according to race and other social factors. Geocoded addresses for residents of other states may result in different patterns of match quality and, consequently, linked exposure measures. We cannot speculate how household addresses including a cancer survivor may differ from households included in research studies of other health conditions.

In conclusion, SAS PROC GEOCODE or ArcGIS geocoders are likely to produce similar results and either tool could be used in collaborative research, but choosing one method over another may depend on project goals and population demographics. For research where reducing misclassification overall and maximizing match rates for rural residents are priorities, scientists may benefit from using ArcGIS software. SAS may have an advantage in terms of convenience for researchers already using SAS for statistical analysis.

## Data Availability

Non-identifiable data in the present study are available upon reasonable request to the authors.

## APPENDIX Supplementary Tables

**Supplementary Table 1:** Demographic characteristics of study participants whose addresses were geocoded.

| Characteristic | WISC |  | Cancer & COVID |  | Total* |  |
| --- | --- | --- | --- | --- | --- | --- |
|  | N | % | N | % | N | % |
| Sex |  |  |  |  |  |  |
| Female | 1554 | 100 | 971 | 57.7 | 2525 | 78.0 |
| Male |  |  | 713 | 42.3 | 713 | 22.0 |
| Age |  |  |  |  |  |  |
| <50 | 12 | 0.8 | 253 | 15.0 | 265 | 8.2 |
| 50-59 | 211 | 13.6 | 316 | 18.8 | 527 | 16.3 |
| 60-69 | 614 | 39.5 | 624 | 37.1 | 1238 | 38.2 |
| ≥70 | 717 | 46.1 | 480 | 28.5 | 1197 | 37.0 |
| Cancer Site |  |  |  |  |  |  |
| Breast | 1554 | 100 | 402 | 23.9 | 1956 | 60.4 |
| Colorectal |  |  | 52 | 3.1 | 52 | 1.6 |
| Prostate |  |  | 215 | 12.8 | 215 | 6.6 |
| Lung |  |  | 84 | 5.0 | 84 | 2.6 |
| Other |  |  | 931 | 55.3 | 931 | 28.8 |
| Race and Ethnicity |  |  |  |  |  |  |
| White, non-Hispanic | 1481 | 95.3 | 1551 | 92.1 | 3032 | 93.6 |
| Other and Multi-racial | 72 | 4.6 | 97 | 5.8 | 169 | 5.2 |
| Education |  |  |  |  |  |  |
| High school degree or less | 634 | 40.8 | 363 | 21.6 | 997 | 30.8 |
| Some college | 418 | 26.9 | 541 | 32.1 | 959 | 29.6 |
| College degree | 338 | 21.7 | 399 | 23.7 | 737 | 22.8 |
| Some graduate school or advanced degree | 164 | 10.6 | 367 | 21.8 | 531 | 16.4 |
| **Total** | 1554 | 100 | 1684 | 100 | 3238 | 100.0 |

\* Numbers may not add up to total N due to missing values, and percentages may not add up to 100% due to missing values.

**Supplementary Table 2:** Publicly available data for geographic linkages to cancer-relevant exposure measures.

| **Measure** | **Geographic Units** | **Year** | **Purpose of Measure and Scoring** | **Citation** |
| --- | --- | --- | --- | --- |
| Yost Index | Block group and census tract | 2014 - 2018 | Measure of socioeconomic status across geographic areas, combining educational level, income, housing, employment<br>Percentile score from 1 (most affluent) to 100 (most deprived) | 10 |
| Environmental Justice (EJ) Index | Census tract | 2022 | The EJ Index ranks each census tract on 36 environmental, social, and health factors and groups them into three overarching modules (Social Vulnerability, Environmental Burden, and Health Vulnerability) and ten different domains. The EJ Index Social-Environmental Ranking (SER) is calculated by combining rankings from the Environmental Burden Module and the Social Vulnerability Module, but not from the Health Vulnerability Module. The SER has a 0-1 range. | 12 |
| Rural-Urban Continuum Codes (RUCCs) | County | 2023 | RUCCs distinguish U.S. metropolitan (metro) counties by the population size of their metro area, and nonmetropolitan (nonmetro) counties by their degree of urbanization and adjacency to a metro area.<br>Metro counties:<br>1 (Counties in metro areas of 1 million population or more)<br>2 (Counties in metro areas of 250,000 to 1 million population)<br>3 (Counties in metro areas of fewer than 250,000 population)<br>Nonmetro counties:<br>4 (Urban population of 20,000 or more, adjacent to a metro area)<br>5 (Urban population of 20,000 or more, not adjacent to a metro area)<br>6 (Urban population of 5,000 to 20,000, adjacent to a metro area)<br>7 (Urban population of 5,000 to 20,000, not adjacent to a metro area)<br>8 (Urban population of fewer than 5,000, adjacent to a metro area)<br>9 (Urban population of fewer than 5,000, not adjacent to a metro area) | 9 |
| Rural-Urban Commuting Area Codes (RUCA) | Census tract | 2010 values revised 7/3/2019 | A measure of urbanization, population density, and daily commuting.<br>1 (Metropolitan area core: primary flow within an urbanized area)<br>2 (Metropolitan area high commuting: primary flow 30% or more to an urbanized area)<br>3 (Metropolitan area low commuting: primary flow 10% to 30% to an urbanized area)<br>4 (Micropolitan area core: primary flow within an urban cluster of 10,000 to 49,999 (large urban core)) | 13 |
|  |  |  | 5 (Micropolitan high commuting: primary flow 30% or more to a large urban core)<br>6 (Micropolitan low commuting: primary flow 10% to 30% to a large urban core)<br>7 (Small town core: primary flow within an urban cluster of 2,500 to 9,999 (small urban core))<br>8 (Small town high commuting: primary flow 30% or more to a small urban core)<br>9 (Small town low commuting: primary flow 10% to 30% to a small urban core)<br>10 (Rural areas: primary flow to a tract outside a urbanized area or urban core)<br>99 (Not coded: Census tract has zero population and no rural-urban identifier information) |  |
| AirToxScreen | Census tract | 2019 | The Air Toxics Screening Assessment (AirToxScreen) is EPA's screening tool that provides communities with information about health risks from air toxics. Specifically, AirToxScreen estimates the cancer risks from breathing air toxics over many years.<br>Columns "A" through "F" provide the geographic location information for that row<br>Column "G" provides the total cancer risk (expressed as risk in 1 million) for that geographic unit | 14 |
| Particulate Matter (PM) 2.5 | latitude/ longitude point within 12 km grid | 2020 | Air quality data based on a Bayesian space-time downscaler model using statistical modeling research in the development of fused space-time predictive surfaces in 12 km grids.<br>Median daily average values from 2020 were calculated from the PM2.5 values in 2020 from the EPA's downscaler model for 12 km grid centroid points in Wisconsin. (PM2.5 describes fine inhalable particles with diameters that are $\leq 2.5$ micrometers averaged over 24 hours) | 17 |
| Distance to TRI Facility | latitude/ longitude point | 2022 | The Toxics Release Inventory (TRI) Release Facility Report, Wisconsin, 2022 for all chemicals and all industries. Including total on-site disposal or other releases, off-site disposal or other releases and total on-and off-site disposal or other releases. Latitude and longitude points are provided for each facility identified in the report. | 18 |

